# Functionality status and drivers of non-functionality of the standardised laboratory analysers in Tanzania mainland: Findings from a National Equipment Harmonisation and Standardisation Mapping Exercise

**DOI:** 10.64898/2026.10.09.26365127

**Authors:** Peter Richard Torokaa, Robert Makala, Joseph Danstan Kitukulu, Esther Mondi Shija, Maria E. Kelly, Godfrey Kwavava, Reuben Sunzu Mkala, Isaac Geofrey Paul, Maige Ngeleja Jidayi, Ritha Yodosi Massawe, David Melville Ocheng, Peter Mashosho Ng’ola, Onna Duuma Panga, Lwitakubi Ferdinand Matata, Louis Nicas Karoli, Alice Kyalo, Alex Sifael Magesa, Majigo Mtebe, Agricola Joachim, Nyambura Moremi

**Author notes:** **Corresponding Author:** Peter Richard Torokaa., Ministry of Health, Dodoma, Tanzania. P.O Box 743, Dodoma.

## Abstract

**Introduction:** Reliable, well-maintained laboratory equipment is essential for timely, accurate and reliable laboratory results for patient care, surveillance, and health system resilience. Tanzania implemented the Standard Medical Laboratory Equipment Guideline (SMLEG-2018) to harmonise procurement and improve efficiency across a tiered laboratory system and network. However, equipment functionality gaps remain a critical challenge for healthcare delivery. This study determined the functionality status of standardised laboratory analysers in Tanzania mainland and identified the drivers of non-functionality.

**Methods:** We conducted a facility-based cross-sectional study to identify the causes of non-functionality of standardised chemistry, haematology, and urine analysers across mainland Tanzania. We assessed 3,044 analysers and used Pareto analysis to identify the primary drivers of non-functionality.

**Results:** Of the 3,044 standardised analysers assessed, 987 (32.4%) chemistry 1,308 (43.0%) haematology, and 749 (24.6%) urine) assessed; 397 (13.0%) were non-functional. Proportionally, urine analysers were the most non-functional (n=176/749, 23.5%), followed by chemistry (n=135/987, 13.7%), and haematology (n=86/1308, 6.6%) analysers, p<0.001. Urine analysers had significantly higher non-functional equipment (23.5%), followed by Chemistry analysers (13.7%), than Haematology analysers (6.6%), p<0.001. Equipment breakdown was the leading cause of non-functionality, accounting for 70.9% (61/86), 70.5% (124/176) and 60.0% (81/135) of the non-functional haematology, urine and chemistry analysers, respectively, among all causes identified for each analyser type.

**Conclusion:** Although the SMLEG 2018 implementation identified broad coverage of functional standardised laboratory equipment, persistent equipment breakdowns of more than 10% highlight a systemic, concerning gap that requires immediate corrective action to sustain high performance. Strengthening preventive maintenance, timely repair services and sustainable equipment lifecycle planning is critical to sustaining quality laboratory services for robust preventive and curative care.

## Introduction

Globally, public health laboratories are recognised as a cornerstone of health systems, providing essential data for disease detection, monitoring, and supporting epidemic preparedness and response (1). Over the last two decades, advancements in laboratory technologies have significantly contributed to improved treatment outcomes and the overall quality of healthcare globally (2). Strengthening laboratory systems and networks is, therefore, a critical global priority, with initiatives led by the World Health Organization (WHO) and partners to improve governance, infrastructure, and workforce capacity (3). Clinical laboratory test reports are essential for disease diagnosis, prognosis, and monitoring, thereby supporting effective treatment and patient management (4).

The WHO African Region’s Regional Strategy on Diagnostic and Laboratory Services (2023– 2032) highlights the importance of harmonising, standardising, and integrating laboratory systems and networks to ensure reliable results and effective disease surveillance (5). In addition, regional assessments have underscored the critical role of laboratories in managing epidemic-prone diseases such as Ebola, Marburg, Cholera, and Measles, which continue to challenge fragile health systems in Africa (6), including Tanzania.

Expanding health care services for communicable diseases (e.g., HIV, Tuberculosis, and Malaria) and non-communicable diseases (e.g., diabetes, cardiovascular and renal diseases) has increased demand for equitable, quality-assured, affordable, and reliable laboratory diagnostics in resource-limited countries. In response to the increased demand for testing services, Tanzania upgraded public laboratories coupled with the introduction of new diagnostic technologies (7). This countermeasure risked further diversifying the existing diagnostic technology architecture and testing platforms, resulting in laboratory networks that are difficult to manage across different healthcare levels.

In line with the objectives of the Tanzania Health Sector Strategic Plan V (2021-2026), different testing methods and procedures must produce quality-assured, comparable, reliable, and consistent results across all healthcare levels of the laboratory systems and networks (8). This can be achieved by standardising and harmonising testing equipment, methodologies, and procedures, coupled with effective nationwide implementation.

Tanzania implemented the Standard Medical Laboratory Equipment Guideline (SMLEG-2018) to harmonise procurement and improve efficiency across a tiered laboratory systems and network. However, equipment functionality gaps remain a critical challenge for healthcare delivery. The guideline highlighted test menus and methodologies by tiered level and discipline to appropriately support clinical services at each level (7). This study therefore aimed to determine the functionality status of laboratory analysers standardised under SMLEG-2018 in Tanzania mainland and to identify the main drivers of their non-functionality.

## Methodology

### Study Design

A facility-based cross-sectional study was conducted to establish the functionality and causes of non-functionality of the standardised chemistry, haematology, and urine analysers across Tanzania mainland.

### Study Area

The evaluation was implemented across 172 of 184 (93.5%) councils in all 26 regions of Tanzania mainland between September and October 2025. The remaining 6.5% of councils were not reached due to access, timing and logistical arrangements.

### Study Population

The study included 3,044 (S1 file) clinical analysers installed across the public health laboratory network under the SMLEG-2018 implementation, comprising chemistry, haematology and urine analysers.

### Data Collection Tools and Procedures

Data collection covered primary and secondary health facilities equipped with standardised analysers under the SMLEG-2018 implementation. The Medical Stores Department (MSD) coordinated facility visits and manages the distribution and installation of laboratory equipment.

Data collectors, including Regional and District Laboratory Technologists, were trained on how to use a validated Excel-based assessment tool prepared by the Ministry of Health (MoH). We used at least one day to assess analyser functionality at each facility. The tool included fields for equipment identification, facility level, functional status, and reasons for non-functionality. The tool further categorised non-functional analysers by cause (e.g., breakdown, reagent shortage, calibration failure). The functionality status was established by direct observation, running a test or internal quality control and the report provided by the laboratory in-charge. The data collector assigned the reason for non-functionality.

### Operational Definitions

#### Functional analyser

This is the analyser able to perform required tests and deliver results as per specifications at the time of assessment.

#### Non-functional analyser

This is the analyser not operational due to breakdown, lack of reagents/consumables, calibration failures, lack of quality control materials, absence of trained laboratory personnel, component failures (e.g., display, printer, data storage), or being classified as beyond repair.

#### Primary and secondary facilities

Defined by level of care under national health system hierarchy. Primary health facilities included (dispensary/health centre) and secondary health facilities included (district/regional hospital).

### Data Analysis

Data were entered, cleaned, and analysed using Epi Info 7, version 7.2.5.0. Frequencies and proportions were generated and were presented in tables and graphs. Pareto analysis, in which causes were ranked by frequency and plotted against their cumulative percentage (80/20 principle), was applied to identify the primary drivers of analyser non-functionality. The relationship of equipment non-functionality was reported with a corresponding p-value of <0.05, reported as significant.

## Results

### Standardised equipment functionality status

Of the 3,044 standardised analysers (S1 file), 987 (32.4%) chemistry 1,308 (43.0%) haematology and 749 (24.6%) urine assessed; 397 (13.0%) were non-functional. The majority of equipment was installed at health centres 2,235 (73.4%), followed by hospitals 717 (23.6%) (**Table 1**).

**Table 1:** Standardised equipment functionality status (N=3,044)

| Characteristics | Frequency | Proportion |
| --- | --- | --- |
| **Distribution of analysers (n=3,044)** |  |  |
| Chemistry analyser | 987 | 32.4 |
| Haematology analyser | 1,308 | 43.0 |
| Urine Analyser | 749 | 24.6 |
| **Non-functionality** |  |  |
| Chemistry analyser (n=987) | 135 | 13.7 |
| Haematology analyser (n=1,308) | 86 | 6.6 |
| Urine analyser (n=749) | 176 | 23.5 |
| **Facility level (n=3,044)** |  |  |
| Dispensary | 92 | 3.0 |
| Health Centre | 2,235 | 73.4 |
| Hospital | 717 | 23.6 |

### Relationship of non-functionality by facility level and equipment type

Proportionally, urine analysers were the most non-functional (n=176/749, 23.5%), followed by chemistry (n=135/987, 13.7%), and haematology (n=86/1308, 6.6%) analysers p<0.001. The non-functionality rate was significantly higher in dispensaries (17.4%), followed by health centres (14.2%), compared with hospitals (8.8%) (p < 0.001). Urine analysers had the highest proportion of non-functional equipment (23.5%), followed by chemistry analysers (13.7%), while haematology analysers had the lowest (6.6%) (p < 0.001) **(Table 2.)**

**Table 2.** Relationship of non-functionality by facility level and equipment type.

| Characteristics | Functionality status |  | p-value |
| --- | --- | --- | --- |
|  | Functional n (%) | Non-functional n (%) |  |
| Facility level |  |  |  |
| Dispensary | 76 (82.6) | 16 (17.4) | <0.001 |
| Health centre | 1917 (85.8) | 318 (14.2) |  |
| Hospital | 654 (91.2) | 63 (8.8) |  |
| Equipment type |  |  |  |
| Urine analyser | 573 (76.5) | 176 (23.5) | <0.001 |
| Chemistry analyser | 852 (86.3) | 135 (13.7) |  |
| Haematology analyser | 1222 (93.4) | 86 (6.6) |  |

### Pareto analysis for drivers of equipment non-functionality

Equipment breakdown was the leading cause of non-functionality, accounting for 70.9% (61/86), 70.5% (124/176) and 60.0% (81/135) of the non-functional haematology, urine and chemistry analysers, respectively. Equipment classified as beyond repair was the second most common cause among haematology (25.6%) and urine (14.2%) analysers, whereas lack of reagents was the second most common cause among chemistry analysers (20.0%) (Table 3). Calibrator failure contributed 13.3% of non-functional chemistry analysers, while the remaining causes each accounted for less than 6% of non-functional analysers (Fig1).

**Table 3.**
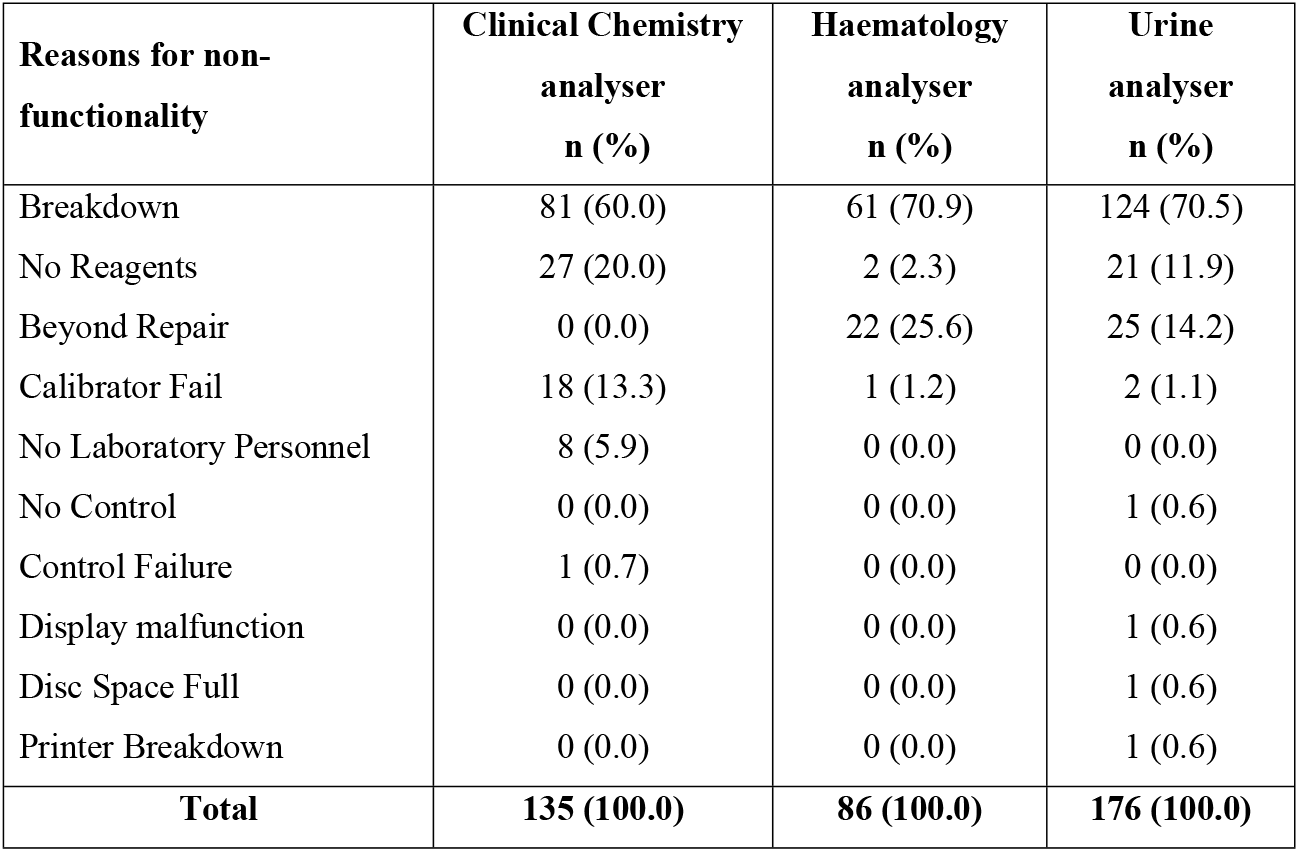
Summary of occurrence of drivers of non-functionality in the standardised laboratory analysers.

| **Reasons for non-functionality** | **Clinical Chemistry analyser**<br>**n (%)** | **Haematology analyser**<br>**n (%)** | **Urine analyser**<br>**n (%)** |
| --- | --- | --- | --- |
| Breakdown | 81 (60.0) | 61 (70.9) | 124 (70.5) |
| No Reagents | 27 (20.0) | 2 (2.3) | 21 (11.9) |
| Beyond Repair | 0 (0.0) | 22 (25.6) | 25 (14.2) |
| Calibrator Fail | 18 (13.3) | 1 (1.2) | 2 (1.1) |
| No Laboratory Personnel | 8 (5.9) | 0 (0.0) | 0 (0.0) |
| No Control | 0 (0.0) | 0 (0.0) | 1 (0.6) |
| Control Failure | 1 (0.7) | 0 (0.0) | 0 (0.0) |
| Display malfunction | 0 (0.0) | 0 (0.0) | 1 (0.6) |
| Disc Space Full | 0 (0.0) | 0 (0.0) | 1 (0.6) |
| Printer Breakdown | 0 (0.0) | 0 (0.0) | 1 (0.6) |
| **Total** | **135 (100.0)** | **86 (100.0)** | **176 (100.0)** |

**Fig 1.**
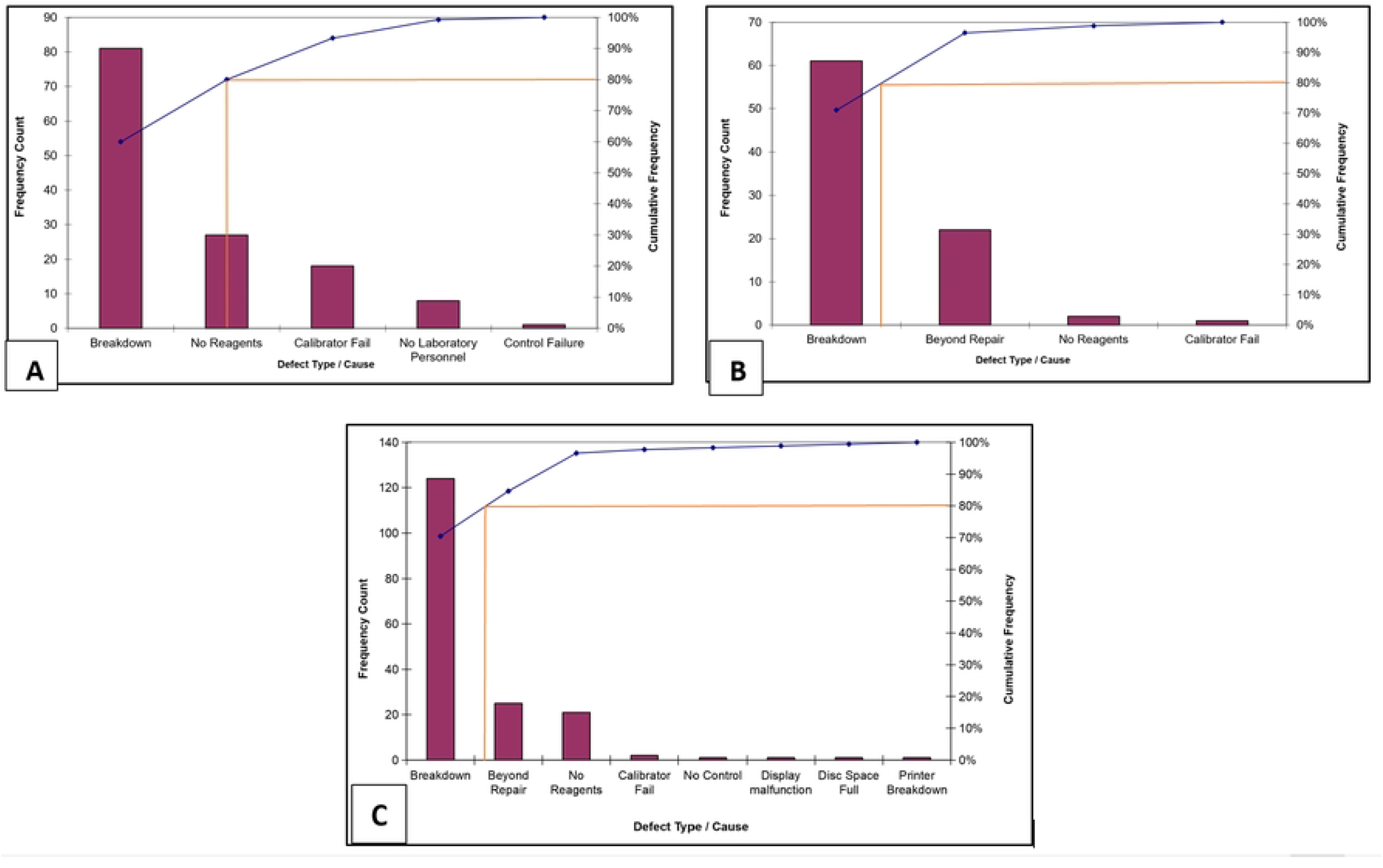
Pareto charts of the drivers of non-functionality among (A) chemistry (n=135), (B) haematology (n=86) and (C) urine (n=176) analysers. Bars show the number of non-functional analysers per cause; the line shows the cumulative percentage, and the horizontal reference line marks the 80% threshold.

## Discussion

This study evaluated Tanzania’s implementation of the SMLEG 2018 and found 13.9% of the assessed analysers were non-functional. Proportionally, urine analysers were the most significantly affected (23.5%), followed by chemistry (13.7%) and haematology (6.6%) analysers, while equipment installed at primary healthcare (Dispensaries and Health centres) had a significantly higher non-functionality rate. Equipment beyond repair was the second most common cause among haematology and urine analysers, whereas lack of reagents was second among chemistry analysers. Reagent stock-outs and calibration failures were other notable causes, particularly for chemistry analysers.

This study found overall equipment non-functionality of 13.0%, 13.7% for chemistry analysers, 6.6% for haematology analysers, and 23.5% for urine analysers. This was lower than in Ethiopia, where a systematic review reported the non-functionality of 28% (7) and 25% for chemistry and haematology analysers (9). Differences in outcomes across countries may reflect variations in infrastructure, funding, and governance. In Tanzania, the relatively low non-functionality rates could be linked to several contextual factors. For example, integrating equipment management into broader laboratory quality improvement initiatives, such as Stepwise Laboratory Quality Improvement Process Towards Accreditation (SLIPTA) programmes, may have contributed, given the number of Tanzanian laboratories enrolled in SLIPTA. Continuous professional training programmes also likely played a role by strengthening staff capacity in preventive maintenance and troubleshooting.

By embedding equipment maintenance and monitoring within national laboratory quality systems, Tanzania aimed to ensure that laboratory personnel were trained not only in technical use but also in preventive maintenance and troubleshooting, which the MSD managed. This emphasis on workforce capability and capacity building and quality assurance frameworks might have contributed in sustaining equipment functionality and maintenance of stronger performance compared to countries without such integrated approaches (10,11). Additionally, the observed low rate of equipment non-functionality may partly be explained by Tanzania’s initiative to harmonise equipment standards across the national laboratory network, which ensured alignment with the first Maputo Declaration (12), and was supported by strong stakeholder collaboration. By addressing procurement inefficiencies, strengthening supply chains, and defining level-specific equipment requirements, Tanzania may have reduced variability and improved the sustainability of laboratory services. These structured interventions likely contributed to the lower non-functionality rates observed compared to settings without similar coordinated frameworks.

The current study found that the leading driver of equipment non-functionality was breakdown (60.0% for chemistry, 70.5% for urine and 70.9% for haematology analysers). This is consistent with a study in Addis Ababa, Ethiopia, which reported that 46% of equipment non-functionality was due to breakdowns (13). In contrast, a study conducted in Jimma Zone, Southwest Ethiopia, identified poor maintenance practices and lack of spare parts as the primary contributors to equipment non-functionality (14). A systematic review by Zamzam et al. highlighted user error and inadequate training as additional factors influencing equipment downtime across healthcare facilities (15). In our setting, breakdowns may reflect a mismatch between equipment workload and limited infrastructural (e.g., power stability) and technical (biomedical engineering and spare-part) support, as well as delayed repairs; however, this study did not capture the underlying causes of breakdown and warrants further investigation. Collectively, these findings indicate that breakdown, often compounded by maintenance gaps, remains the dominant driver of equipment non-functionality across settings.

These findings have practical implications. First, because breakdown accounted for most non-functional analysers, strengthen planned preventive maintenance through enforceable service-level agreements with suppliers, adequate regional and zonal biomedical engineering capacity, and routine monitoring of equipment uptime. Second, formally decommission and replace analysers classified as beyond repair through lifecycle-based budgeting to avoid prolonged service gaps. Third, reagent stock-outs and calibration failures, particularly for chemistry analysers, highlight the need to align reagent and calibrator forecasting and supply with the installed equipment base and test volumes.

This study has some limitations. First, only 172 of 184 (93.5%) councils were reached; 3,044 (94.9%) of the 3,207 installed standardised analysers were assessed, and the status of the remaining 163 (5.1%) analysers could not be determined. Second, functionality was assessed at a single point in time, so the study could not determine the duration and frequency of downtime. Third, the study recorded only one primary reason per non-functional analyser, so it did not capture underlying causes of breakdown (e.g., equipment age, power supply, user handling, availability of spare parts, and service contracts). However, the large sample size of standardised laboratory equipment was a major strength of this study, as it enhanced visibility and transparency regarding the success of laboratory equipment harmonisation and standardisation in Tanzania.

## Conclusion

Although SMLEG-2018 implementation has achieved wide coverage of functional standardised laboratory equipment, the overall non-functionality rate of 13.0%, driven predominantly by equipment breakdown, highlights systemic gaps that require immediate corrective actions for sustained high performance. Strengthening preventive maintenance, timely repair services, reliable reagent supply and sustainable equipment lifecycle planning is critical to sustaining quality laboratory services for robust preventive and curative care.

## Data Availability

All data are in the manuscript and the supporting information files

## Ethical considerations

The study evaluated the implementation of SMLEG-2018 within the MoH framework for routine services without direct access to patient information; therefore, MoH granted permission to publish the findings and a waiver through a letter (Ref. no. MP.76232/06), dated 19^th^ May 2026. Before analysis, data was anonymised to ensure the confidentiality of facilities, equipment, and testing platforms.

### Authors contributions

**Conceptualization:** Peter Richard Torokaa, Nyambura Moremi.

**Data curation:** Peter Richard Torokaa, Joseph Danstan Kitukulu, Esther Mondi Shija and Peter Ngo’llah Mashosho, Godfrey Kwavava and Robert Makala, Nyambura Moremi.

**Formal analysis:** Peter Richard Torokaa, Mtebe Majigo

**Methodology:** Peter Richard Torokaa

**Supervision:** Nyambura Moremi.

**Validation:** Joseph Danstan Kitukulu, Esther Mondi Shija and Peter Ngo’llah Mashosho, Godfrey Kwavava and Robert Makala

**Writing-original draft:** Peter Richard Torokaa, Nyambura Moremi, David Melville Ocheng, Maria E. Kelly, Mtebe Majigo, Agricola Joachim, Alice Kyalo

**Writing – review & editing:** Peter Richard Torokaa, Robert Makala, Joseph Danstan Kitukulu, Esther Mondi Shija, Maria E. Kelly, Godfrey Kwavava, Reuben Sunzu Mkala, Isaac Geofrey Paul, Maige Ngeleja Jidayi, Ritha Yodosi Massawe, David Melville Ocheng, Peter Ngo’llah Mashosho, Onna Duuma Panga, Lwitakubi Ferdinand Matata, Louis Nicas Karoli, Alice Kyalo, Alex Sifael Magesa, Majigo Mtebe, Agricola Joachim, Nyambura Moremi

### Funding

The study did not receive any specific funding.

### Data availability statement

All data are in the manuscript and the supporting information files

## Declarations

### Informed Consent

Not Applicable.

### Consent for publication

Not applicable

### Competing interests

The authors declare that they have no competing interests.

---

**S1 file. Standardized equipment dataset**

## Notes

### Competing Interest Statement

The authors have declared that no competing interests exist.

### Author Declarations

The study evaluated the implementation of SMLEG-2018 within the MoH framework for routine services without direct access to patient information therefore, MoH granted permission to publish the findings and a waiver through a letter (Ref. no. MP.76232/06), dated 19th May 2026. Before analysis, data was anonymised to ensure the confidentiality of facilities, equipment, and testing platforms.

